# Adherence to CDC Antimicrobial Stewardship Core Elements and Barriers to stewardship practices among Healthcare Workers at a Tertiary Care Hospital Uttarakhand, India

**DOI:** 10.64898/2026.03.26.26349469

**Authors:** Manisha, Kirti Kumari, Komal Meena, Kusum, Mamta Pilania, Mahima Kashyap, Komal Mahala, Manisha Bhakar, Neetu Kataria, Vijayshree, Prasan K Panda, Maneesh Sharma

## Abstract

**Background:** Antimicrobial resistance (AMR) is a growing global health concern driven largely by inappropriate antimicrobial use. Antimicrobial stewardship programs (ASPs), guided by the Centers for Disease Control and Prevention (CDC) core elements, are essential for optimizing antimicrobial use. However, adherence to these practices and the barriers faced by healthcare workers remain inadequately explored, particularly in resource-limited settings.

**Objective:** To assess adherence to the CDC antimicrobial stewardship checklist and identify barriers affecting stewardship practices among healthcare workers at a tertiary care hospital in Uttarakhand, India.

**Methods:** A quantitative cross-sectional descriptive study was conducted among 355 healthcare workers, including nursing officers and physicians. Data were collected using a sociodemographic questionnaire, the CDC antimicrobial stewardship checklist, and a self-structured barrier assessment tool (test–retest reliability r = 0.78). Descriptive and inferential statistics were applied using SPSS version 23.0, with a significance level set at p ≤ 0.05.

**Results:** The overall adherence to the CDC antimicrobial stewardship checklist was 52.3%, indicating moderate compliance. Higher adherence was observed in action-oriented interventions, while lower adherence was noted in domains such as accountability, pharmacy expertise, reporting, and education. Major barriers identified included lack of antimicrobial supply (89.0%), shortage of key personnel (88.5%), delays in laboratory reports (85.1%), lack of training (83.9%), and inadequate administrative support (79.2%). Significant associations were found between perceived barriers and factors such as working area, designation, qualification, and work experience (p < 0.05), whereas age and gender showed no significant association.

**Conclusion:** Adherence to antimicrobial stewardship practices was moderate, with notable gaps in organizational and educational components. Multiple systemic, resource-related, and behavioral barriers hinder effective implementation. Targeted interventions focusing on strengthening infrastructure, workforce capacity, training, and administrative support are essential to improve stewardship practices in tertiary care settings.

## Introduction

Antimicrobial resistance (AMR) represents an escalating global health challenge that threatens the effective prevention and treatment of infectious diseases^1^. The inappropriate, irrational, and excessive use of antimicrobial agents in healthcare settings is a major contributing factor to the emergence and spread of resistant microorganisms^2,3^. Healthcare institutions play a pivotal role in combating AMR through the implementation of antimicrobial stewardship programs (ASPs), which are designed to optimize antimicrobial prescribing, improve patient outcomes, minimize adverse effects, and reduce the development of resistance^4,5^. In this context, the Centers for Disease Control and Prevention (CDC) has established core elements for hospital antimicrobial stewardship programs, providing a structured framework for implementation and evaluation^6^.

Despite the increasing recognition of ASPs, adherence to standardized stewardship practices among healthcare workers remains inconsistent, particularly in resource-constrained settings such as India^7-9^. Factors such as limited infrastructure, workforce shortages, inadequate training, and lack of institutional support may hinder effective implementation^10^. Furthermore, there is a paucity of evidence exploring both adherence to stewardship frameworks and the barriers faced by frontline healthcare providers in tertiary care settings. Understanding these factors is essential for designing targeted interventions to strengthen stewardship efforts. Therefore, the present study was undertaken to assess adherence to the CDC antimicrobial stewardship checklist and to identify barriers affecting stewardship practices among healthcare workers at a tertiary care hospital in Uttarakhand.

## Methods

A quantitative cross-sectional descriptive study was conducted at a tertiary care teaching hospital in Uttarakhand, India. The study population comprised 355 healthcare workers, including nursing officers and physicians, working in selected clinical areas. The sample size was calculated using OpenEpi, considering a population size of 2000, an expected adherence proportion of 50%, a precision of 5%, a design effect of 1, and a 10% allowance for non-response. Participants were recruited using a convenience sampling technique based on eligibility criteria, which included willingness to participate and availability during the data collection period.

Data were collected using three instruments: a sociodemographic questionnaire, the CDC antimicrobial stewardship checklist, and a self-structured questionnaire to assess barriers to antimicrobial stewardship practices. The barrier questionnaire consisted of 14 items and demonstrated acceptable reliability, with a test– retest reliability coefficient of r = 0.78. Content validity was ensured through expert evaluation by professionals in infection control, microbiology, and nursing. Data collection was carried out between October 2024 and February 2025 after obtaining informed consent from participants, ensuring confidentiality and anonymity throughout the process.

The collected data were entered into Microsoft Excel and analyzed using SPSS version 23.0. Descriptive statistics, including frequency, percentage, mean, and standard deviation, were used to summarize the data. Inferential statistical tests, including Chi-square test, Mann–Whitney U test, Kruskal–Wallis test, and Spearman’s correlation, were applied to assess associations between variables. A p-value of ≤ 0.05 was considered statistically significant. Ethical approval for the study was obtained from the Institutional Ethics Committee of the tertiary care hospital.

## Results

The sociodemographic profile of 355 healthcare workers depicted mean age of 29.26 ± 3.08 years. Slightly more than half of the participants were male (50.8%). Nursing officers constituted the largest proportion of the sample, followed by junior residents. A considerable proportion of participants held a Bachelor of Science in Nursing degree, and the mean duration of work experience was approximately 57 months. Notably, only a minority of participants had prior exposure to antimicrobial stewardship activities, with 17.5% having participated in AMS committees and 32.8% having attended related training programs. The table presents the adherence to various components of the CDC Antimicrobial Stewardship Program (ASP) checklist across seven domains among healthcare facilities.

The table 1 shows variable adherence to the CDC Antimicrobial Stewardship Program components across seven domains. In the leadership domain, compliance ranged from 46.2% to 66.5%, with better support for dedicated stewardship time but weaker integration into quality improvement activities. Accountability and pharmacy expertise were suboptimal, with less than half of facilities having designated leaders (45.4%) or trained pharmacists (∼45%). Intervention-related practices showed relatively higher adherence, particularly for common infections such as urinary tract infections (78.3%) and sepsis (73.5%), although structured strategies like antibiotic time-outs were less frequently implemented (47.3%). Tracking mechanisms were inconsistently applied, with moderate monitoring of documentation (60.0%) but low participation in national surveillance systems (38.6%–41%). Similarly, reporting and feedback practices were inadequate, with less than half of facilities sharing data with prescribers. Educational activities were also limited, reported by only 43–47% of facilities. Overall, the findings indicate moderate to low adherence across most stewardship domains, with comparatively better performance in clinical interventions and significant gaps in leadership, accountability, tracking, reporting, and education.The overall adherence to the CDC antimicrobial stewardship checklist was found to be 52.3%, indicating moderate compliance among healthcare workers (Fig 1). Among the various domains of the checklist, the highest adherence was observed in action-oriented interventions aimed at improving antibiotic use. In contrast, lower adherence levels were reported in domains such as accountability, pharmacy expertise, reporting systems, and educational initiatives, highlighting gaps in institutional and organizational components of stewardship.

**Table 1.** Frequency distribution of adherence to CDC checklist (N=355)

| S. N | Elements | Yes<br>f (%) | No<br>f (%) |
| --- | --- | --- | --- |
| **Domain 1** | **Hospital Leadership Commitment** |  |  |
| A1 | Does facility leadership provide stewardship program leader(s) dedicated time to manage the program and conduct daily stewardship interventions? | 236 (66.5) | 119 (33.5) |
| A2 | Does facility leadership provide stewardship program leader(s) with resources (e.g. IT support, training) to effectively operate the program? | 202 (56.9) | 153 (43.1) |
| A3 | Does your antibiotic stewardship program have a senior executive that serves as a point of contact or “champion” to help ensure the program has resources and support to accomplish its mission | 182 (51.3) | 173 (48.7) |
| A4 | Do stewardship program leader(s) have regularly scheduled meetings with facility leadership and/or the hospital board to report and discuss stewardship activities, resources and outcomes? | 173 (48.7) | 182 (51.3) |
| A5 | Does your facility leadership ensure that staff from key support departments and groups have sufficient time to contribute to stewardship activities? | 177 (49.9) | 178 (50.1) |
| A6 | Does facility leadership ensure that antibiotic stewardship activities are integrated into other quality improvement and patient safety efforts, such as sepsis management and diagnostic stewardship? | 164 (46.2) | 191 (53.8) |
| A7 | Does facility leadership support enrolment and reporting into the National Healthcare Safety Network (NHSN) Antimicrobial Use and Resistance (AUR) Module, including any necessary IT support? | 179 (49.3) | 180 (50.7) |
| **Domain 2** | **Accountability** |  |  |
| B1 | Does your facility have a leader or co-leaders responsible for program management and outcomes of stewardship activities? | 161 (45.4) | 194 (54.6) |
| Ba | If a non-physician is the leader of the program, does the facility have a designated physician who can serve as a point of contact and support for the non-physician leader? | 134 (37.7) | 221 (62.3) |
| **Domain 3** | **Pharmacy Expertise** |  |  |
| C1 | Does your facility have a pharmacist(s) responsible for leading implementation efforts to improve antibiotic use? | 162 (45.6) | 192 (54.1) |
| C2 | Does your pharmacist(s) leading implementation efforts have specific training and/or experience in antibiotic stewardship? | 161 (45.4) | 194 (54.6) |
| **Domain 4** | **Action: Implement Interventions to Improve Antibiotic Use** |  |  |
| D1 | Does your facility perform prospective audit and feedback for specific antibiotic agents? | 193 (54.4) | 162 (45.6) |
| D2 | Does your facility perform preauthorization for specific antibiotic agents? | 174 (49.0) | 181 (51.0) |
| D3 | Does your facility have facility-specific treatment recommendations, based on national guidelines and local pathogen susceptibilities, to assist with antibiotic selection for common clinical conditions | 170 (47.9) | 184 (51.8) |
| D4a | Does your facility have specific interventions (e.g., ensuring correct discharge duration of therapy) to ensure optimal use of antibiotics for treating the most common infections in most hospitals?<br>a. Community-acquired pneumonia | 202 (56.9) | 153 (43.1) |
| D4b | Urinary tract infections | 278 (78.3) | 77 (21.7) |
| D4c | Skin and soft tissue infections | 238 (67.0) | 117 (33.0) |
| D5a | . Does your facility have specific interventions in place to ensure optimal use of antibiotics in the following situations?<br>a. Sepsis | 261 (73.5) | 94 (26.5) |
| D5b | Staphylococcus aureus infection | 251 (70.7) | 104 (29.3) |
| D5c | Stopping unnecessary antibiotic(s) in new cases of Clostridium difficile infection (CDI) | 219 (61.7) | 136 (38.3) |
| D5d | Culture-proven invasive (e.g., blood stream) infection | 235 (66.2) | 120 (33.8) |
| D5e | Review of planned outpatient parenteral antibiotic therapy (OPAT) | 222 (62.5) | 133 (37.5) |
| D6 | Does your facility have a policy that requires prescribers to document in the medical record or during order entry a dose, duration, and indication for all antibiotic prescriptions? | 241 (67.9) | 114 (32.1) |
| D7 | Does your facility have a formal procedure for all prescribers to conduct daily reviews of antibiotic selection until a definitive diagnosis and treatment duration are established (i.e. time out)? | 168 (47.3) | 187 (52.7) |
| **Domain 5** | **Tracking Antibiotic Use and Outcomes** |  |  |
| E1 | Does your antibiotic stewardship program track antibiotic use by submitting to the National Healthcare Safety Network (NHSN) Antimicrobial Use (AU) Option? | 137 (38.6) | 218 (61.4) |
| E2 | Does your antibiotic stewardship program monitor prospective audit and feedback interventions by tracking the types of interventions and acceptance of recommendations? | 133 (37.5) | 222 (62.5) |
| E3 | Does your antibiotic stewardship program monitor preauthorization interventions by tracking which agents are being requested for which conditions? | 148 (41.7) | 207 (58.3) |
| E4 | Does your stewardship program monitor adherence to facility specific treatment recommendations? | 196 (55.2) | 159 (44.8) |
| E5 | Does your stewardship program monitor adherence to a documentation policy (dose, duration, and indication)? | 213 (60.0) | 142 (40.0) |
| E6 | Does your antibiotic stewardship program monitor the performance of antibiotic timeouts to see how often these are being done and if opportunities to improve use are being acted on during timeouts? | 181 (51.0) | 174 (49.0) |
| E7 | Does your antibiotic stewardship program routinely perform medication use evaluations to assess courses of therapy for select antibiotics and/or infections to identify opportunities to improve use? | 167 (47.0) | 188 (53.0) |
| E8 | Does your antibiotic stewardship program assess how often patients are discharged on the correct antibiotics for the recommended duration? | 196 (55.2) | 159 (44.8) |
| E9 | Does your antibiotic stewardship program track antibiotic resistance by submitting to the NHSN Antimicrobial Resistance (AR) Option? | 148 (41.7) | 207 (58.3) |
| E10 | Does your antibiotic stewardship program track CDI in context of antibiotic use? | 141 (39.7) | 214 (60.3) |
| E11 | Does your facility produce an anti-bio gram (cumulative antibiotic susceptibility report)? | 171 (48.2) | 184 (51.8) |
| **Domain 6 Reporting Antibiotic Use and Outcomes** |  |  |  |
| F1 | Does your antibiotic stewardship program share facility and/or individual prescriber-specific reports on antibiotic use with prescribers? | 172 (48.5) | 183 (51.5) |
| F2 | Does your antibiotic stewardship program report adherence to treatment recommendations to prescribers (e.g., results from medication use evaluations, etc.)? | 163 (45.9) | 192 (54.1) |
| F3 | Has your facility distributed a current anti bio gram to prescribers? | 164 (46.2) | 191 (53.8) |
| **Domain 7 Education** |  |  |  |
| G1 | Does your stewardship program provide education to prescribers and other relevant staff on optimal prescribing, adverse reactions from antibiotics, and antibiotic resistance? | 167 (47.0) | 181 (53.0) |
| G2 | Does your stewardship program provide education to prescribers as part of the prospective audit and feedback process (sometimes called “handshake stewardship”)? | 154 (43.4) | 201 (56.6) |

**Figure.1:**
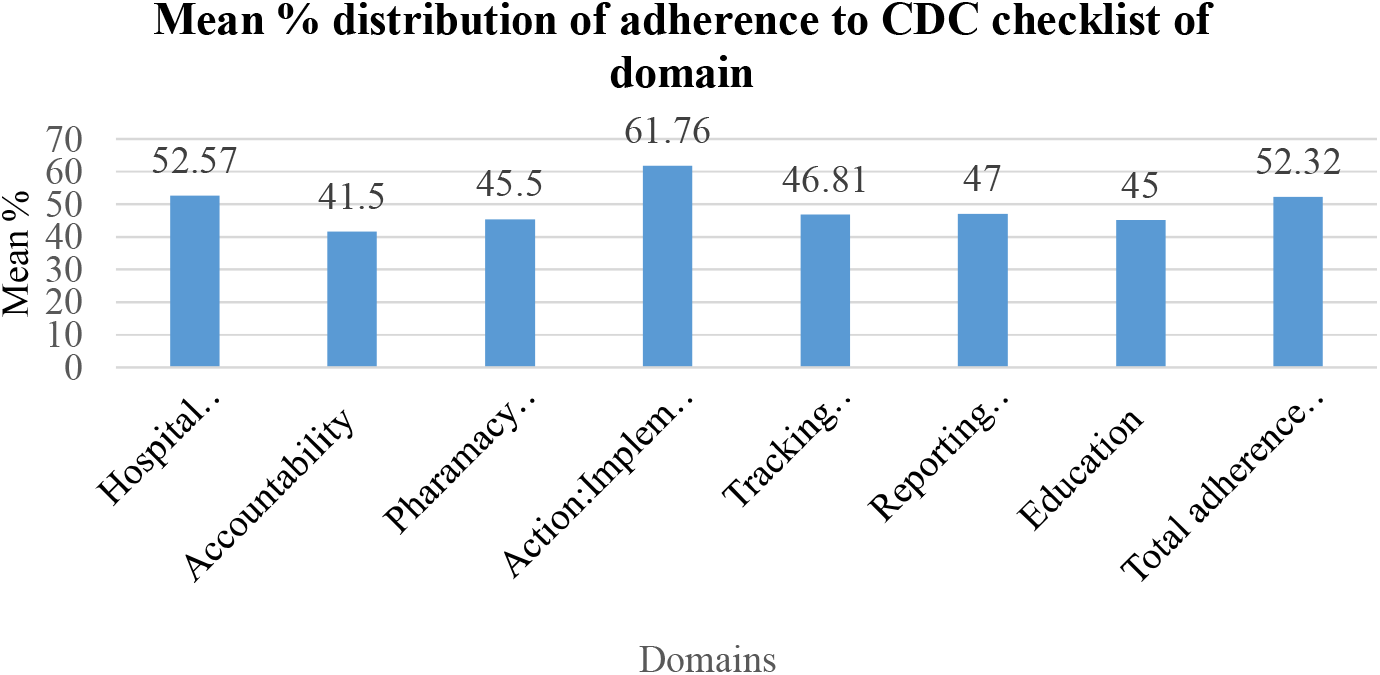
Mean % distribution of adherence to CDC checklist of domain.

The table 2 highlights the perceived barriers to antimicrobial stewardship practices among healthcare workers, with a majority reporting multiple challenges. The most commonly identified barriers were lack of antimicrobial supply (89.0%), shortage of key personnel such as infectious disease specialists and pharmacists (88.5%), and delays in laboratory reports, particularly antibiotic sensitivity testing (85.1%). Other prominent barriers included lack of training and orientation (83.9%), poor administrative reinforcement (82.8%), and inadequate human resources (82.0%). Additionally, issues such as poor communication among healthcare workers (80.6%), financial constraints (80.3%), and lack of administrative support (79.2%) were widely reported. Structural and system-level barriers like absence of reliable antibiograms (78.9%), time constraints (78.6%), and lack of national guidelines (78.0%) were also noted. Furthermore, reluctance among healthcare workers to modify prescriptions due to fear of patient deterioration (73.2%) and limited IT infrastructure (69.9%) contributed to suboptimal implementation of stewardship practices. Overall, the findings indicate that antimicrobial stewardship is hindered by a combination of resource limitations, organizational challenges, and behavioral factors.

**Table 2.**
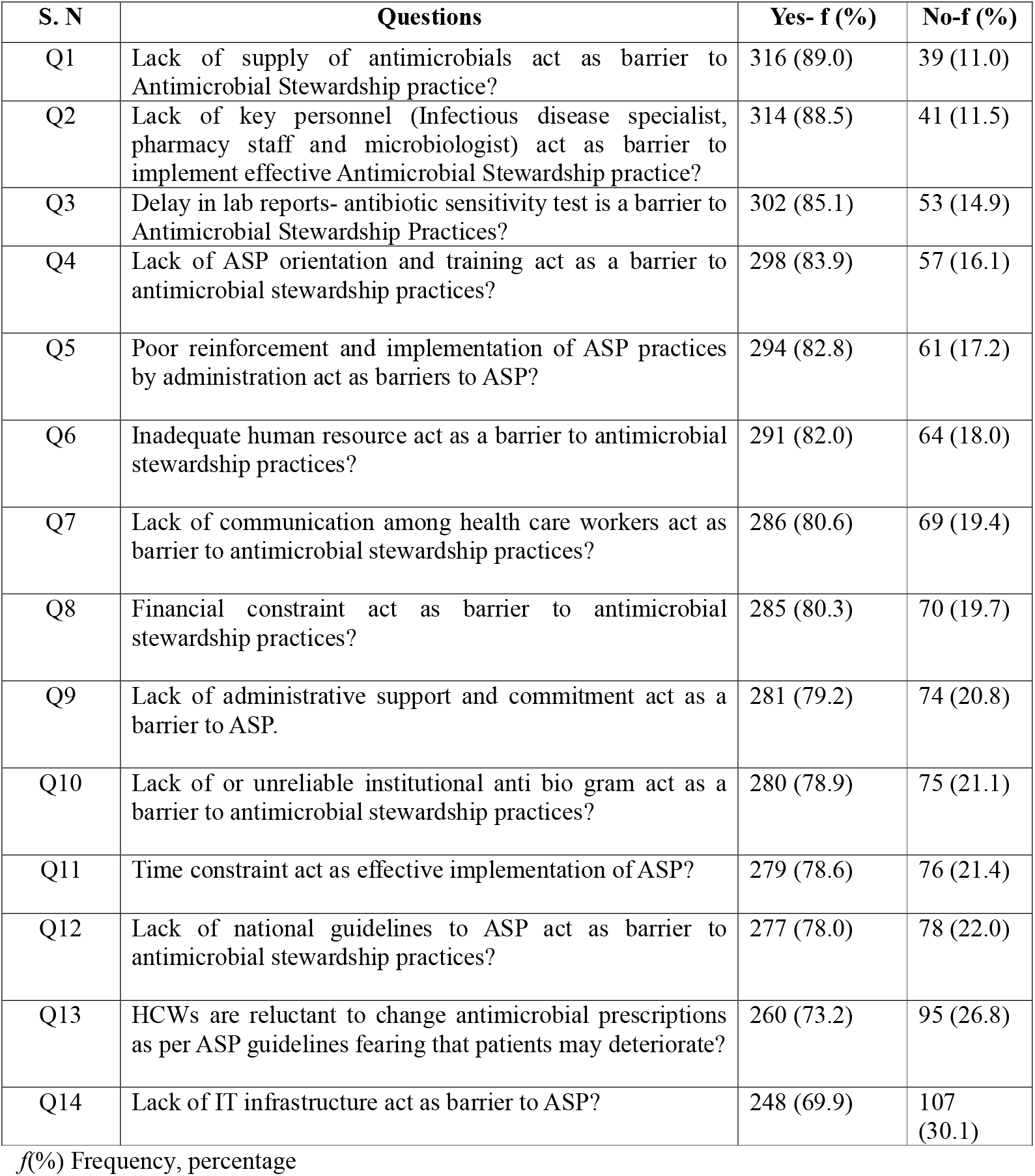
Frequency distribution of barriers to antimicrobial stewardship practice. (N=355)

The table 3 depicts the association between perceived barriers to antimicrobial stewardship practices and selected socio-demographic variables. Overall, most barriers did not show a statistically significant association with age, gender, research involvement, or committee membership. However, several factors demonstrated significant associations. Working area was significantly associated with multiple barriers, including lack of antimicrobial supply, lack of institutional antibiogram, lack of ASP orientation, lack of national guidelines, financial constraints, lack of communication, lack of administrative support, lack of IT infrastructure, and time constraints (p < 0.05). Designation and qualification were also significantly associated with key barriers such as lack of institutional antibiogram, lack of ASP orientation, lack of national guidelines, financial constraints, lack of communication, and lack of administrative support. Additionally, total work experience showed significant association with barriers like lack of institutional antibiogm, lack of ASP orientation, lack of national guidelines, financial constraints, lack of communication, and lack of administrative support (p < 0.05). Participation in workshops was significantly associated with lack of national guidelines and poor reinforcement of ASP practices. These findings suggest that organizational and professional characteristics, particularly working area, designation, qualification, and experience, play a crucial role in influencing perceived barriers to antimicrobial stewardship practices.

**Table 3.**
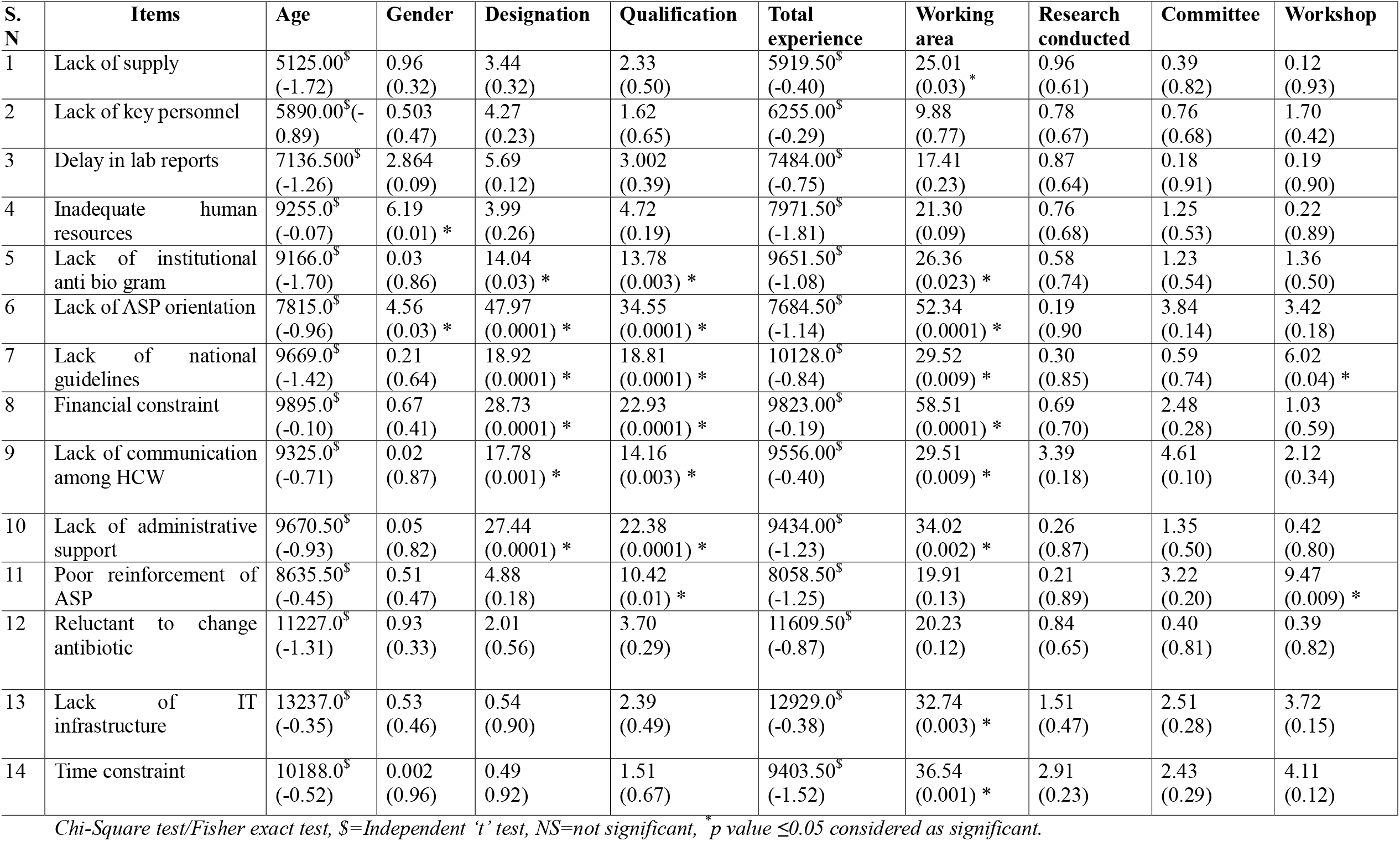
Association of Barrier with selected socio-demographic Variables (N=355)

| S. N | Items | Age | Gender | Designation | Qualification | Total experience | Working area | Research conducted | Committee | Workshop |
| --- | --- | --- | --- | --- | --- | --- | --- | --- | --- | --- |
| 1 | Lack of supply | 5125.00<sup>\$</sup><br>(-1.72) | 0.96<br>(0.32) | 3.44<br>(0.32) | 2.33<br>(0.50) | 5919.50<sup>\$</sup><br>(-0.40) | 25.01<br>(0.03) * | 0.96<br>(0.61) | 0.39<br>(0.82) | 0.12<br>(0.93) |
| 2 | Lack of key personnel | 5890.00<sup>\$</sup>(-0.89) | 0.503<br>(0.47) | 4.27<br>(0.23) | 1.62<br>(0.65) | 6255.00<sup>\$</sup><br>(-0.29) | 9.88<br>(0.77) | 0.78<br>(0.67) | 0.76<br>(0.68) | 1.70<br>(0.42) |
| 3 | Delay in lab reports | 7136.500<sup>\$</sup><br>(-1.26) | 2.864<br>(0.09) | 5.69<br>(0.12) | 3.002<br>(0.39) | 7484.00<sup>\$</sup><br>(-0.75) | 17.41<br>(0.23) | 0.87<br>(0.64) | 0.18<br>(0.91) | 0.19<br>(0.90) |
| 4 | Inadequate human resources | 9255.0<sup>\$</sup><br>(-0.07) | 6.19<br>(0.01) * | 3.99<br>(0.26) | 4.72<br>(0.19) | 7971.50<sup>\$</sup><br>(-1.81) | 21.30<br>(0.09) | 0.76<br>(0.68) | 1.25<br>(0.53) | 0.22<br>(0.89) |
| 5 | Lack of institutional anti bio gram | 9166.0<sup>\$</sup><br>(-1.70) | 0.03<br>(0.86) | 14.04<br>(0.03) * | 13.78<br>(0.003) * | 9651.50<sup>\$</sup><br>(-1.08) | 26.36<br>(0.023) * | 0.58<br>(0.74) | 1.23<br>(0.54) | 1.36<br>(0.50) |
| 6 | Lack of ASP orientation | 7815.0<sup>\$</sup><br>(-0.96) | 4.56<br>(0.03) * | 47.97<br>(0.0001) * | 34.55<br>(0.0001) * | 7684.50<sup>\$</sup><br>(-1.14) | 52.34<br>(0.0001) * | 0.19<br>(0.90) | 3.84<br>(0.14) | 3.42<br>(0.18) |
| 7 | Lack of national guidelines | 9669.0<sup>\$</sup><br>(-1.42) | 0.21<br>(0.64) | 18.92<br>(0.0001) * | 18.81<br>(0.0001) * | 10128.0<sup>\$</sup><br>(-0.84) | 29.52<br>(0.009) * | 0.30<br>(0.85) | 0.59<br>(0.74) | 6.02<br>(0.04) * |
| 8 | Financial constraint | 9895.0<sup>\$</sup><br>(-0.10) | 0.67<br>(0.41) | 28.73<br>(0.0001) * | 22.93<br>(0.0001) * | 9823.00<sup>\$</sup><br>(-0.19) | 58.51<br>(0.0001) * | 0.69<br>(0.70) | 2.48<br>(0.28) | 1.03<br>(0.59) |
| 9 | Lack of communication among HCW | 9325.0<sup>\$</sup><br>(-0.71) | 0.02<br>(0.87) | 17.78<br>(0.001) * | 14.16<br>(0.003) * | 9556.00<sup>\$</sup><br>(-0.40) | 29.51<br>(0.009) * | 3.39<br>(0.18) | 4.61<br>(0.10) | 2.12<br>(0.34) |
| 10 | Lack of administrative support | 9670.50<sup>\$</sup><br>(-0.93) | 0.05<br>(0.82) | 27.44<br>(0.0001) * | 22.38<br>(0.0001) * | 9434.00<sup>\$</sup><br>(-1.23) | 34.02<br>(0.002) * | 0.26<br>(0.87) | 1.35<br>(0.50) | 0.42<br>(0.80) |
| 11 | Poor reinforcement of ASP | 8635.50<sup>\$</sup><br>(-0.45) | 0.51<br>(0.47) | 4.88<br>(0.18) | 10.42<br>(0.01) * | 8058.50<sup>\$</sup><br>(-1.25) | 19.91<br>(0.13) | 0.21<br>(0.89) | 3.22<br>(0.20) | 9.47<br>(0.009) * |
| 12 | Reluctant to change antibiotic | 11227.0<sup>\$</sup><br>(-1.31) | 0.93<br>(0.33) | 2.01<br>(0.56) | 3.70<br>(0.29) | 11609.50<sup>\$</sup><br>(-0.87) | 20.23<br>(0.12) | 0.84<br>(0.65) | 0.40<br>(0.81) | 0.39<br>(0.82) |
| 13 | Lack of IT infrastructure | 13237.0<sup>\$</sup><br>(-0.35) | 0.53<br>(0.46) | 0.54<br>(0.90) | 2.39<br>(0.49) | 12929.0<sup>\$</sup><br>(-0.38) | 32.74<br>(0.003) * | 1.51<br>(0.47) | 2.51<br>(0.28) | 3.72<br>(0.15) |
| 14 | Time constraint | 10188.0<sup>\$</sup><br>(-0.52) | 0.002<br>(0.96) | 0.49<br>(0.92) | 1.51<br>(0.67) | 9403.50<sup>\$</sup><br>(-1.52) | 36.54<br>(0.001) * | 2.91<br>(0.23) | 2.43<br>(0.29) | 4.11<br>(0.12) |

Chi-Square test/Fisher exact test, \$=Independent 't' test, NS=not significant, \* $p$ value $\leq 0.05$ considered as significant.

## Discussion

The findings of the present study indicate moderate adherence to the Centers for Disease Control and Prevention antimicrobial stewardship core elements among healthcare workers in a tertiary care setting. Similar levels of partial implementation have been reported in previous studies, where clinical interventions such as antibiotic optimization were more frequently practiced than administrative and structural components. Simlarly Pollack LA et al.^11^ reported that while action and tracking elements were relatively well established, domains such as accountability and reporting remained underdeveloped in many hospitals. This aligns with the current findings, where deficiencies were evident in accountability, pharmacy expertise, reporting mechanisms, and educational initiatives.

The high prevalence of reported barriers in the present study is consistent with findings from resource-constrained settings. Studies conducted by Raka L et al.^12^ and Charani E et al.^13^ have highlighted that shortages of trained personnel, limited microbiology support, and inadequate institutional backing are major challenges in implementing antimicrobial stewardship programs. Delays in diagnostic services and lack of structured training programs, as identified in the present study, the similar findings were reported by Rajendran R et al.^14^ where healthcare systems often face infrastructural and logistical constraints.

The observed significant association between adherence and factors such as designation, qualification, and participation in stewardship committees is supported by earlier research. Abbo LM et al.^15^ demonstrated that healthcare professionals with specialized training and direct involvement in stewardship initiatives exhibited higher adherence to antimicrobial guidelines. Likewise, Kakkar AK et al.^16^ emphasized the role of continuous professional education and interdisciplinary collaboration in improving antimicrobial prescribing practices. These findings reinforce the importance of structured capacity-building programs and active engagement of healthcare workers in stewardship committees.

Furthermore, the need for a comprehensive and multifaceted approach identified in this study is strongly supported by global recommendations. The World Health Organization and CDC both emphasize leadership commitment, accountability frameworks, workforce training, and robust surveillance systems as essential components of effective antimicrobial stewardship programs. Studies across different healthcare settings have consistently shown that institutions with strong administrative support and integrated stewardship policies achieve better outcomes in antimicrobial use and resistance control.

Overall, the findings of the present study are in concordance with existing literature, highlighting that while progress has been made in implementing antimicrobial stewardship practices, significant gaps remain in organizational support, infrastructure, and education. Addressing these barriers through policy strengthening, investment in laboratory services, and continuous professional development is critical for optimizing antimicrobial use and combating the growing threat of antimicrobial resistance.

## Conclusion

The present study demonstrates moderate adherence to CDC antimicrobial stewardship practices among healthcare workers in a tertiary care hospital in Uttarakhand, accompanied by a high burden of systemic and organizational barriers. These findings highlight the urgent need for strengthening institutional support, enhancing workforce capacity, improving diagnostic and laboratory services, and promoting structured training programs. A comprehensive and sustained effort at the organizational and policy level is essential to improve antimicrobial stewardship practices and effectively address the challenge of antimicrobial resistance.

## Data Availability

All data produced in the present work are contained in the manuscript

## Declarations

### Ethics approval and consent to participate

Approved by the Institutional Ethics Committee of AIIMS Rishikesh. Written informed consent was obtained from all participants.

### Author contributions

Conceptualization, data collection, analysis, and manuscript preparation were performed by the authors.

## Acknowledgements

The authors acknowledge the healthcare workers of AIIMS Rishikesh for their participation.

